# Trajectories of Hypoxemia & Respiratory System Mechanics of COVID-19 ARDS in the NorthCARDS dataset

**DOI:** 10.1101/2021.01.26.21250492

**Authors:** Daniel Jafari, Amir Gandomi, Alex Makhnevich, Michael Qiu, Daniel M Rolston, Eric P Gottesman, Adey Tsegaye, Paul H Mayo, Molly E Stewart, Meng Zhang, Negin Hajizadeh

## Abstract

**Rationale:** The preliminary reports of COVID Acute Respiratory Distress Syndrome (COVIDARDS) suggest the existence of a subset of patients with higher lung compliance despite profound hypoxemia. Understanding heterogeneity seen in patients with COVIDARDS and comparing to non-COVIDARDS may inform tailored treatments.

**Objectives:** To describe the trajectories of hypoxemia and respiratory compliance in COVIDARDS and associations with outcomes.

**Methods:** A multidisciplinary team of frontline clinicians and data scientists created the Northwell COVIDARDS dataset (NorthCARDS) leveraging over 11,542 COVID-19 hospital admissions. Data was summarized to describe differences based on clinically meaningful categories of lung compliance, and compared to non-COVIDARDS reports. A sophisticated method of extrapolating PaO2 from SpO2, as well estimating FiO2 from non invasive oxygen delivery devices were utilized to create meaningful trends of derived PaO2 to FiO2 (P/F).

**Measurements and Main Results:** Of the 1595 COVIDARDS patients in the NorthCARDS dataset, there were 538 (34·6%) who had very low lung compliance (<20ml/cmH2O), 982 (63·2%) with low-normal compliance (20-50ml/cmH2O), and 34 (2·2%) with high lung compliance (>50ml/cmH2O). The very low compliance group had double the median time to intubation compared to the low-normal group (107 hours(IQR 26·3, 238·3) vs. 37·9 hours(IQR 4·8, 90·7)). Oxygenation trends have improved in all groups after a nadir immediately post intubation. The P/F ratio improved from a mean of 109 to 155, with the very low compliance group showing a smaller improvement compared to low compliance group. The derived P/F trends closely correlated with blood gas analysis driven P/F trends, except immediately post intubation were the trends diverge as illustrated in the image. Overall, 67·5% (n=1049) of the patients died during the hospitalization. In comparison to non-COVIDARDS reports, there were less patients in the high compliance category (2.2%vs.12%, compliance ≥50mL/cmH20), and more patients with P/F ≤ 150 (57·8% vs. 45.6%). No correlation was apparent between lung compliance and P/F ratio. The Oxygenation Index was similar, (11·12(SD 5·67)vs.12·8(SD 10·8)).

**Conclusions:** Heterogeneity in lung compliance is seen in COVIDARDS, without apparent correlation to degree of hypoxemia. Notably, time to intubation was greater in the very low lung compliance category. Understanding ARDS patient heterogeneity must include consideration of treatment patterns in addition to trajectories of change in patient-level data and demographics.

## INTRODUCTION

A subset of patients with COVID-19 deteriorate despite supportive measures, requiring invasive mechanical ventilation for acute respiratory failure and the acute respiratory distress syndrome (ARDS).^2^Controversy has existed regarding the differences between COVID-related ARDS (COVIDARDS) versus other causes of ARDS.^3–5^ For example, there appear to be a subset of patients with higher lung compliance despite profound hypoxemia.^6^ The cohorts of COVIDARDS patients reported in the literature have been limited by sample size and have not included data on pulmonary mechanics. Herein, we describe the development of the NorthCARDS dataset which includes data on trajectories of illness, response to treatments and lung compliance for COVID-19 patients who were mechanically ventilated at one of the 12 acute care hospitals within the Northwell Health System. We then describe differences in demographics and treatments as well as trajectories of lung compliance and hypoxemia over the hospital course for patients with very low versus low-normal lung compliance. This work sets the stage for further data analytics among patients with COVIDARDS to better characterize phenogroups using readily available data elements from electronic health records.

## METHODS

### Patients, Study Design and Data Collection

This is a retrospective study of intubated and mechanically ventilated patients with ARDS and COVID-19 who were admitted to one of 12 acute care hospitals within the Northwell Health System during the height of the pandemic in NYC (March 1 - April 30, 2020). Northwell Health is the largest academic health system in New York, serving approximately 11 million people. Discharge disposition was available until Sept 30^th^, 2020 for all patients in the cohort. Within the Northwell Health COVID-19 Research Consortium, the Northwell ARDS Research Collaborative was formed by a multi-disciplinary group of clinical providers and research scientists (data scientists, biostatisticians and clinical trialists) to work on the creation of the NorthCARDS dataset.

All patients admitted to one of the 12 hospitals within the Northwell Health system during the time period of March 1 through April 30, 2020 were screened. Inclusion criteria were: Age > 18, COVID-19 polymerase chain reaction (PCR) test positive during the hospitalization, treatment with invasive mechanical ventilation, and PaO_2_: Fraction of inspired Oxygen (FiO_2_) ratio, (referred to as P/F), of less than 300 for 2 consecutive measurements during the hospital admission. The requirement for bilaterality of infiltrates as per the Berlin ARDS definition was confirmed based on a random sample of one hundred patients who were reviewed for radiographic findings of bilateral pulmonary involvement based on attending radiologist read of chest x-rays or CT scan. Surgical patients, identified by the presence of a brief operative note within 24 hours of intubation time (Ti) were excluded unless the mechanical ventilation was for a post-operative state rather than for the procedure alone.

Features relevant for understanding patients’ lung mechanics were extracted from the electronic health records of COVIDARDS patients. All available laboratory values, medications and oxygen supplementation concentration and mode as well as pulse oximetry results (SpO_2_) were recorded.

This study was considered by Northwell Health Institutional Review Board as minimal-risk using data collected for routine clinical practice and waived the requirement for informed consent.

### Data Definitions and Assumptions

Several data assumptions needed to be made to structure the data. These included which fields contained the most valid and reliable data, and how best to handle missing data. For transparency, we outline assumptions for data structuring below and how we tested these assumptions. The Northwell ARDS Research Collaborative discussed each assumption to ensure that they reflected the clinical practice of providers caring for patients and their data entry in to the electronic health record. Further details are provided in the Supplement.

#### Oxygen Delivery Method, Concentration, and Degree of Hypoxemia

The FiO_2_ delivered was calculated based on the following formula: for nasal cannula or non-rebreather face mask, each liter of oxygen flow added 0·04 to 0·21 (room air), with a maximum of 6 liters per minute for nasal cannula and 15 liters per minute for non-rebreather mask. In the instances where the delivery method was not recorded in the electronic medical record, the previous recorded method was presumed to be have been continued, until change in flow rate or delivery method was noted. To be able to accurately map hypoxemia prior to intubation, we used both arterial blood gas data on partial pressure of oxygen (PaO_2_) and peripherally measured oxygen saturation (SpO_2_). We calculated SpO_2_:FiO_2_ ratios as well as PaO_2_:FiO_2_ ratios over time for each patient across their entire hospital stay. For separate analyses we converted SpO_2_:FiO_2_ to PaO_2_:FiO_2_ ratios (‘derived P/F’) to obtain an estimated trajectory of PaO_2_ over time^7^ (derived P/F = ([SpO2:FiO2]-64)/0.84). The assumption that derived P/F would have parallel trends compared to ABG based P/F was visually tested (Figure 4).

#### Respiratory System Compliance

We used both static compliance (change in lung volume per unit change in pressure in the *absence of flow)* using the plateau pressure recorded in the electronic medical record, (Tidal Volume/ [Plateau Pressure – Peak End Expiratory Pressure (PEEP]); and dynamic compliance using the Peak Inspiratory Pressure (PIP) (change in lung volume per unit change in pressure in the *presence of flow)*, (Tidal Volume/ [PIP – PEEP]) when patients were deeply sedated/paralyzed as described below. We only included values obtained at the time of full patient sedation, which was defined as the administration of intermittent bolus or continuous infusions of paralytics and difference in patient respiratory rate and set respiratory rate <2 breaths/minute (Figure S·7). We made the assumption that patients would not have a significant component of airway resistance for most COVID-19 respiratory failure patients in the early stage of disease (no more than a difference of 5-7 cmH2O between PIP and Plateau pressures), and that therefore this added pressure due to flow would have a minimal contribution to overall measured compliance. This assumption was tested by visualizing the difference between static and dynamic compliance seen over time (Figure S·3).

### Outcomes measured

In addition to establishing the NorthCARDS dataset, we sought to explore whether there were different phenogroups of COVIDARDS. The primary outcome was the number of patients in categories of lung compliance on the first day of ARDS, and the characteristics seen descriptively in each category. The secondary outcome was hospital mortality and discharge location. Ventilator parameters and respiratory mechanics were reported for each group of pre-defined compliance. While oxygenation, pulmonary mechanics and therapeutics were censored on June 23^rd^, the hospital disposition data was available through the end of September.

### Statistical analyses

Descriptive statistics included proportions for categorical variables, and mean (standard deviation) and median (interquartile range) for continuous variables. We used the independent-samples t-test, proportions z-test, and/or Mood’s median test to compare very low and low-normal compliance categories, and two-sided p-value <0·05 as the threshold of statistical significance. The data was analyzed using Python 3·7 and several libraries including pandas, numpy, matplotlib, scipy, nltk, and re. Because the size of our dataset could lead to finding statistically significant associations without clinical significance, each outcome was reviewed for clinical significance by the clinicians in the Northwell ARDS Collaborative and results are discussed in the context of pathophysiological validity and other investigators’ results.

## RESULTS

We identified 3176 patients who were admitted between March 1 and April 30, 2020 to one of the Northwell Health System hospitals, and who were mechanically ventilated. Of these, 2020 patients were COVID-19 PCR positive and 1554 met inclusion criteria with reliable lung compliance data (Figure 1). Data for patients who were excluded are presented in the supplement (Table S1). Discharge disposition for index hospitalization was available for all patients except two patients.

**Figure 1.**
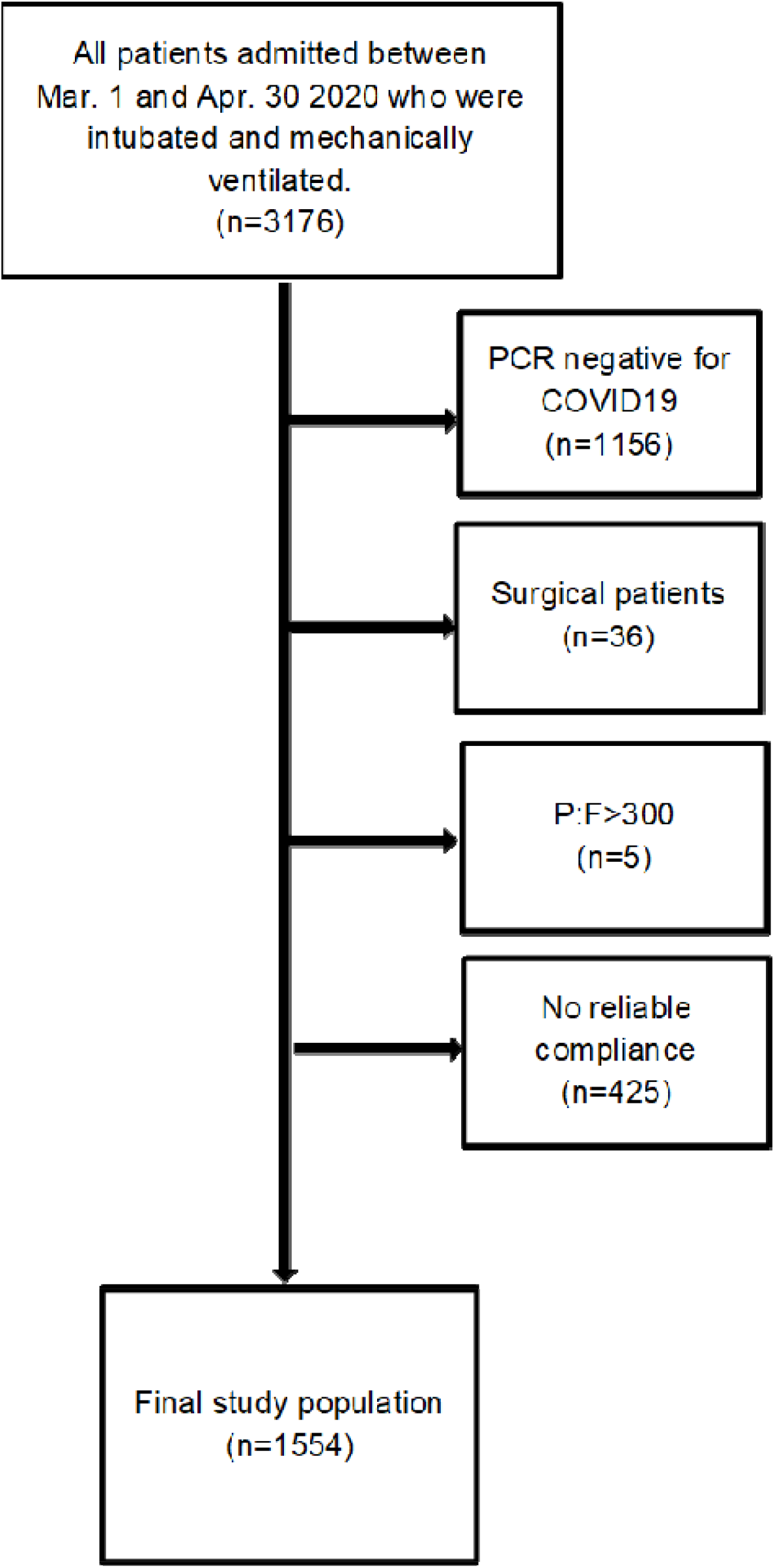
Flowchart of study population and exclusions

### Lung Compliance categories

The average lung compliance for the whole cohort was 24·44 mL/cm H2O (SD 11·69). Frequencies per decile of compliance are presented in Figure 2. Based on clinical observations, the Northwell ARDS Collaborative chose to categorize the cohort into three categories: very low compliance (<20 mL/cm H2O); low-normal (20-50 mL/cm H2O) and high (> 50 mL/cm H2O) measured by the dynamic compliance over the first 24 hours of intubation in the setting of paralytics or deep sedation. There were 538 (34·6%) patients with very low compliance; 982 (63· 2%) with low-normal compliance, and 34 (2·2%) with high compliance. Given the very small sample size in the higher compliance category, comparators of prevalence and exploratory statistical testing is limited to the very low versus low-normal compliance groups. The average median difference between static and dynamic compliance overall was 6·41 mL/cm H2O (IQR 3·16, 11·42, n=1053). For the very low compliance group median difference was 4·60 mL/cm H2O (IQR 2·05, 8·10, n=429); and for the low-normal group 7·89 mL/cm H2O (IQR 4·19, 12·64, n=610).

**Figure 2.**
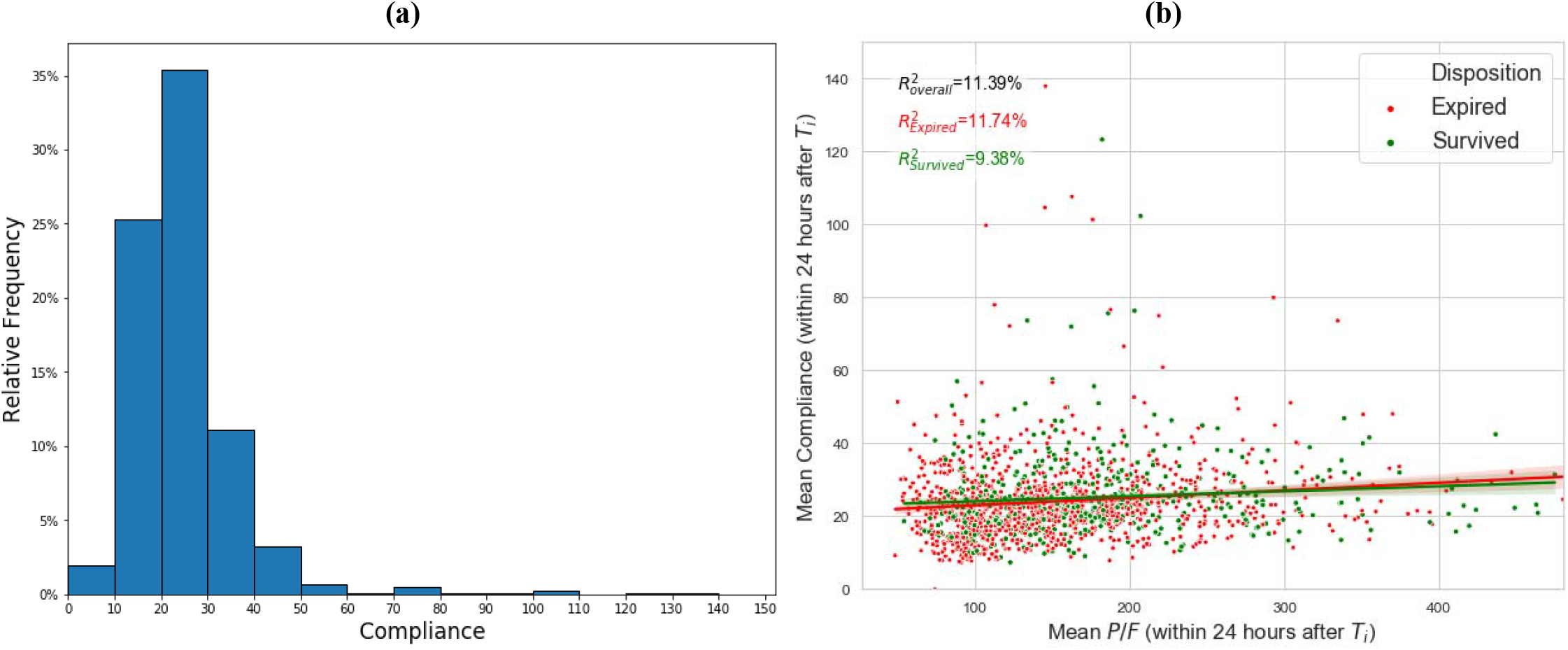
**(a)** Frequency of lung compliance seen in the first 24 hours of mechanical ventilation for the entire cohort with reliable compliance data (N=1554) by decile, and **(b)** Correlation between Compliance and P/F from ABGs in the first 24 hours of intubation.

### COVIDARDS Demographics

Patient demographics are detailed in Table 1. Overall, average age was 65 years, 32% were female, 35% were white, and the average Charlson comorbidity index was 4·9 (SD 3·3) (corresponding to a roughly 52% estimated 1-year survival).^8^ The MEWS score was also high (4·1, SD 1·9) (corresponding to a roughly 12·7% chance of ICU admission or death within 60 days).^9^ There was a greater percentage of females in the very low compliance category (43·7%, vs. 24·9%, low-normal; and 29·4%,high), and more were non-white/multi-racial. The most common comorbidity was hypertension (65·1%, n=1012) and diabetes (43·4%, n=675). The overall cohort included 16% (n=251) with BMI indicating extreme obesity (BMI >40).

**Table 1.** Demographics stratified by high/normal/low compliance

|  | All patients with reliable compliance (n=1554) | Patients with very low compliance <20 ml/cm H<sub>2</sub>O (n=538; 34.6%) | Patients with low-normal compliance 20–50 ml/cm H<sub>2</sub>O (n=982; 63.2%) | Patients with high compliance >50 ml/cm H<sub>2</sub>O (n=34; 2.2%) | p-value (very low vs. low-normal compliance) |
| --- | --- | --- | --- | --- | --- |
| **Age median, IQR** | 65.0 [56.0,73.0] | 64.0 [56.0,73.0] | 65.5 [56.0,74.0] | 69.0 [60.2,73.0] | 0.025 |
| **Female n (%)** | 490 (31.5) | 235 (43.7) | 245 (24.9) | 10 (29.4) | < 0.001 |
| **Race n (%)** |  |  |  |  |  |
| **Black/African American** | 290 (18.7) | 113 (21.0) | 172 (17.5) | 5 (14.7) | 0.096 |
| **Asian** | 154 (9.9) | 70 (13.0) | 82 (8.4) | 2 (5.9) | 0.004 |
| **Other/Multiracial** | 484 (31.1) | 193 (35.9) | 272 (27.7) | 17 (50.0) | < 0.001 |
| **Unknown** | 81 (5.2) | 25 (4.6) | 54 (5.5) | 2 (5.9) | 0.47 |
| **White** | 545 (35.1) | 137 (25.5) | 400 (40.7) | 8 (23.5) | < 0.001 |
| **Comorbidities (Charlson Index) mean (SD)** | 4.9 (3.3) | 4.7 (3.2) | 5.0 (3.3) | 4.4 (2.2) | 0.094 |
| **MEWS on admission mean (SD)** | 4.1 (1.9) | 4.2 (1.8) | 4.1 (1.9) | 3.9 (1.6) | 0.301 |
| **Chronic Lung Disease n (%)** | 102 (6.6) | 33 (6.1) | 67 (6.8) | 2 (5.9) | 0.60 |
| **Diabetes n (%)** | 675 (43.4) | 211 (39.2) | 378 (38.5) | 14 (41.2) | 0.28 |
| **HTN n (%)** | 1012 (65.1) | 349 (64.9) | 645 (65.7) | 18 (52.9) | 0.75 |
| **CHF n (%)** | 114 (7.3) | 20 (3.7) | 53 (5.4) | 1 (2.9) | 0.14 |
| **CAD n (%)** | 208 (13.4) | 57 (10.6) | 147 (15) | 4 (11.8) | 0.017 |
| **CKD n (%)** | 228 (14.7) | 67 (12.5) | 158 (16.1) | 3 (8.8) | 0.056 |
| **ESRD n (%)** | 85 (5.5) | 24 (4.5) | 60 (6.1) | 1 (2.9) | 0.178 |
| **Positive Smoking n (%)** | 195 (15.5) | 63 (13.9) | 131 (16.7) | 1 (4.2) | 0.18 |
| **Malignancy n (%)** | 147 (9.5) | 51 (9.5) | 94 (9.6) | 2 (5.9) | 0.037 |
| **BMI on admission mean (SD)** | 31.0 (7.8) | 30.5 (7.4) | 31.4 (8.1) | 29.5 (4.3) | 0.025 |
| **BMI Categories n (%)** |  |  |  |  |  |
| **BMI <18 underweight** | 10 (0.6) | 7 (1.3) | 3 (0.3) | 0 | 0.0217 |
| **BMI 18 – <30 normal–overweight** | 740 (47.6) | 267 (49.6) | 457 (46.5) | 16 (47.1) | 0.25 |
| **BMI 30 – <40 obese** | 553 (35.6) | 185 (34.4) | 353 (35.9) | 15 (44.1) | 0.54 |
| **BMI >=40 extremely obese** | 251 (16.2) | 79 (14.7) | 169 (17.2) | 3 (8.8) | 0.20 |

### Interventions/treatments

Almost all patients (89%, n=1383) received hydroxychloroquine, 62% (n=963) received azithromycin, 82% (n=1278) received steroids, 52% (n=815) received paralytics, and 49% (n=769) were proned. IL-1 and IL-6 inhibitors were given to 30% (n=475) of the patients, while 7% received convalescent plasma (n=109). During the first 48 hours, 83·5% (n=449) received at least one vasopressor in the very low compliance category, compared to 77·2% (n=758) in the low-normal group. (Table 2)

**Table 2.** Interventions stratified by compliance groups

| During the entire hospital stay | All patients with reliable compliance<br>N=1554 | Patients with very low compliance<br><20 ml/cm H<sub>2</sub>O<br>(n=538; 34.6%) | Patients with low-normal compliance<br>20–50 ml/cm H<sub>2</sub>O<br>(n=982; 63.2%) | Patients with high compliance<br>>50 ml/cm H<sub>2</sub>O<br>(n=34; 2.2%) | p-value<br>(very low vs. low-normal compliance) |
| --- | --- | --- | --- | --- | --- |
| **Steroids\*** n (%) | 1278 (82.1) | 467 (86.8) | 780 (79.4) | 29 (85.3) | < 0.001 |
| **Hydroxychloroquine** n (%) | 1383 (89) | 459 (85.3) | 893 (90.9) | 31 (91.2) | < 0.001 |
| **Azithromycin** n (%) | 963(62) | 269 (50.0) | 672 (68.4) | 22 (64.7) | < 0.001 |
| **IL–1 or IL–6 inhibitor\*\*** n (%) | 475 (30.6) | 190 (35.3) | 274 (27.9) | 11 (32.4) | 0.003 |
| **Remdesivir** n (%) | 12 (0.7) | 5 (0.9) | 7 (0.7) | 0 | 0.650 |
| **Convalescent plasma** n (%) | 109 (7) | 46 (8.6) | 62 (6.3) | 1 (2.9) | 0.100 |
| **Proning** | 769 (49.4) | 319 (59.3) | 436 (44.4) | 14 (41.2) | < 0.001 |
| **Paralytics\*\*\*** | 815 (52.4) | 313 (58.2) | 487 (49.6) | 14 (41.2) | 0.001 |
| **Vasopressors (y/n) in first 48hrs**<br>n (%) | 1,232 (79.2) | 449 (83.5) | 758 (77.2) | 25 (73.5) | 0.004 |
| **Inotropes (y/n) at any time**<br>count n (%) | 64 (4.1) | 28 (5.2) | 0 (0.0) | 36 (3.6%) | < 0.001 |
| **Pre-intubation O<sub>2</sub> supplementation n (%) \*\*\*\*** |  |  |  |  |  |
| **NRB** | 1196 (77) | 426 (79.2) | 743 (75.7) | 27 (79.4) | 0.119 |
| **NRB+NC** | 41(2.6) | 18 (3.3) | 23 (2.3) | 0 | 0.248 |
| **NC** | 82 (5.3) | 21 (3.9) | 59 (6.0) | 2 (5.9) | 0.079 |
| **HFNC** | 33 (2.1) | 13 (2.4) | 20 (2) | 0 | 0.628 |
| **NIV (BiPAP/CPAP)** | 51 (3.3) | 23 (4.3) | 27 (2.7) | 1(2.9) | 0.111 |
| **Venturi** | 15 (1) | 2 (0.4) | 13 (1.3) | 0 | 0.072 |
| **Other\*\*\*\*\*** | 136 (8.8) | 35 (6.5) | 97 (9.9) | 4 (11.8) | 0.026 |
| **Median hours from hospital presentation to intubation (IQR)** | 50.8 [7.5,123.7] | 107.0 [26.3,238.3] | 37.9 [4.8,90.7] | 29.4 [1.0,79.4] | < 0.001 |
| * dexamethasone, hydrocortisone, methylprednisolone, prednisone, prednisolone<br>** Anikinra, Tocilizumab, Sarilumab<br>***Rocuronium, Vecuronium, Cisatracurium<br>**** NRB= Nonrebreather Mask; NC- Nasal Canula; HFNC= High Flow Nasal Canula; NIV = Non-Invasive Mechanical Ventilation; Venturi = Venturi Mask<br>***** Other included room air; BVM; tracheostomy collar; simple face; bag mask; ambubag; T-piece; king airway; |  |  |  |  |  |

### Time to Intubation

On average, COVIDARDS patients were intubated within 50·8 hours (IQR 7·5,123·7) from the time of admission. Patients in the very low compliance group had the longest time between admission and intubation, 107 hours (IQR 26·3, 238·3), compared to 37·9 hours (IQR 4·8, 90·7) in the low-normal compliance group. Prior to intubation, 77% (n=1196) of patients were receiving oxygen supplementation via non rebreather masks, with 2·1% (n=33) on HFNC, and 3·3% (n=51) on NIV, which reflects infection control practices at the time discouraging NIV use. (Table 2)

### P/F ratios and blood gas results

The average blood gas pH in the 24 hour period before intubation was 7·29 (SD 0·14), and PaCO_2_ was 51·4 mmHg (SD 19·31). (Table 3) Patients in the very low lung compliance category had higher levels of PaCO_2_ and lower mean arterial pH. ABG was not performed in 70·8% cases during the 12 hours prior to intubation. The overall mean derived P/F ratio in the 12 hours prior to intubation was 95 (SD 85), which was lowest for those in the high compliance group (P/F 66, SD 33) (Figure 5). When including PEEP in the calculation of P/F ratio, the P/FPEEP (PFP)^10^ also appeared lowest for those in the highest compliance category. There was no correlation between P/F and compliance. (Figure 2)

**Table 3.**
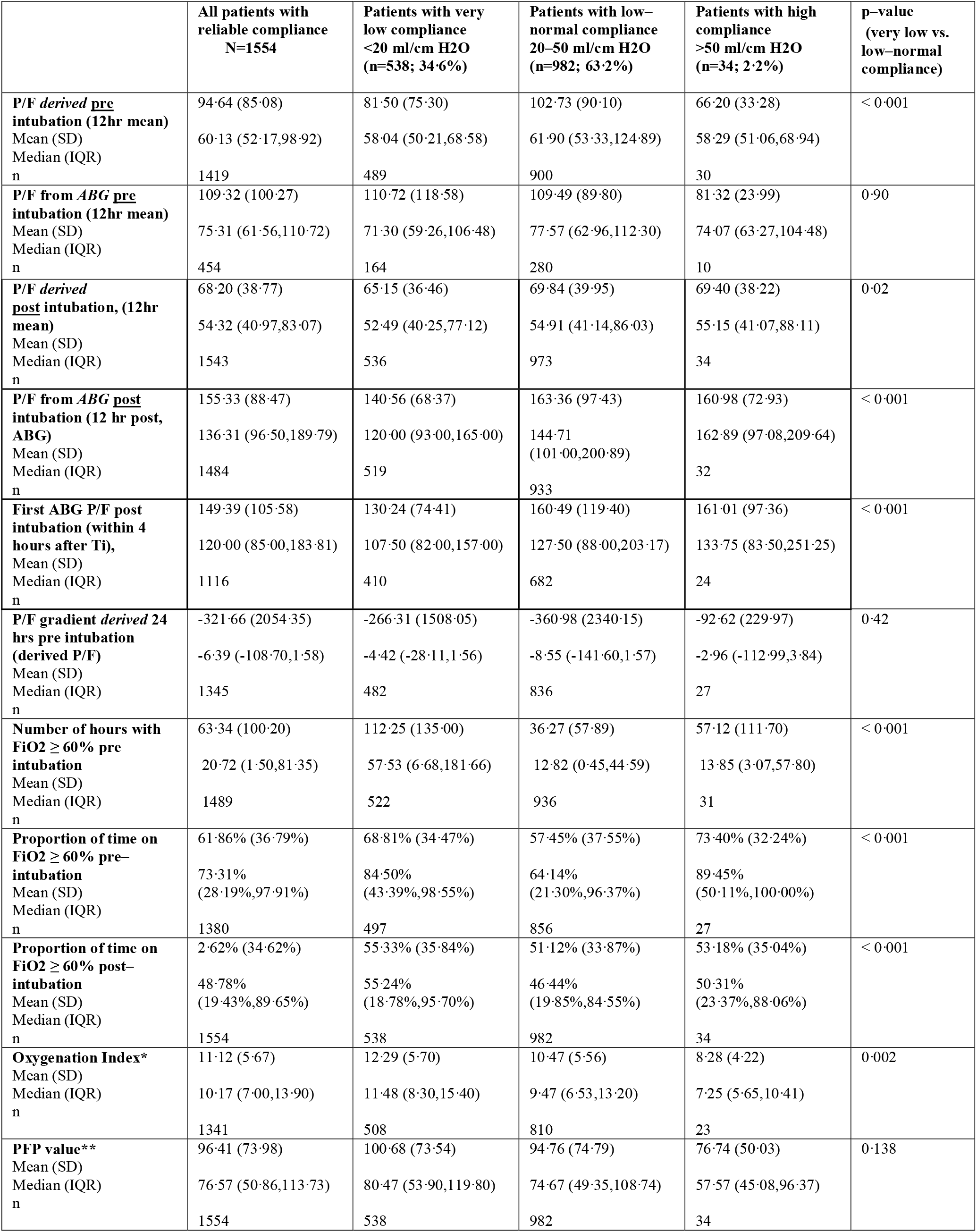
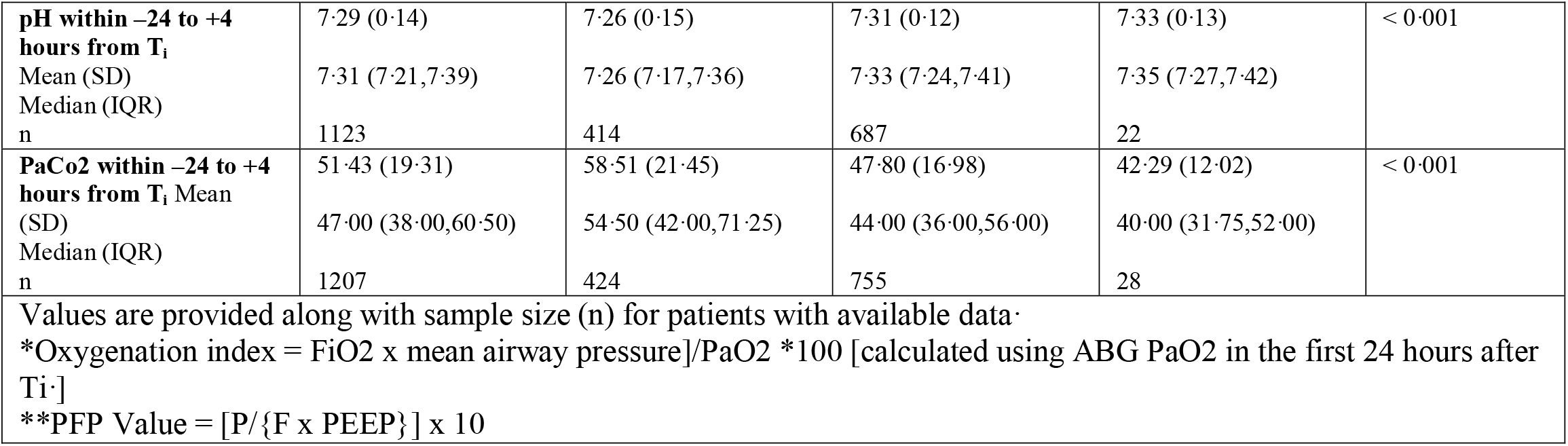
Oxygenation trends and duration of ventilation by compliance group

**Figure 3.**
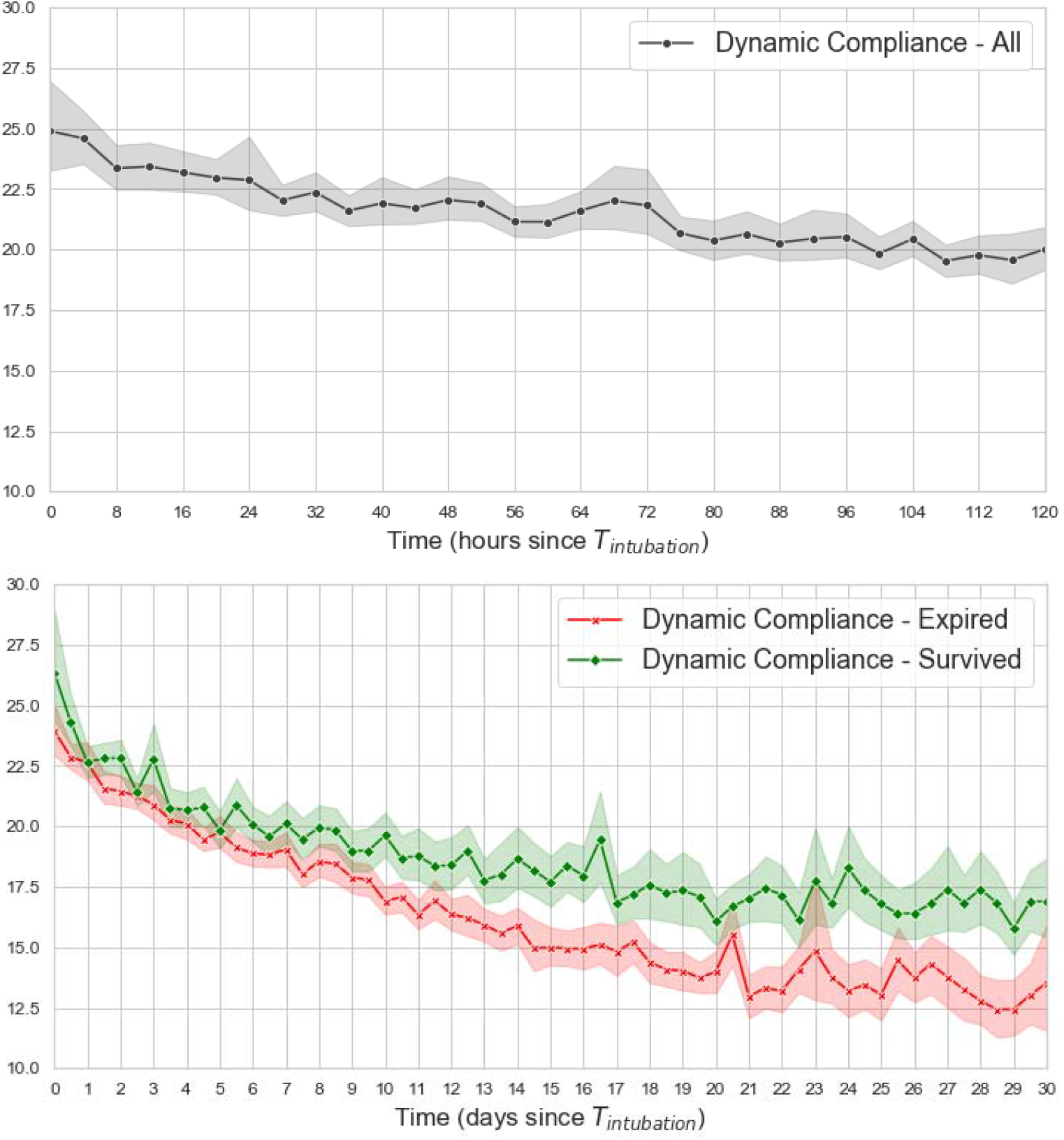
Trends in compliance for the total cohort [N= 1554] for the first 5 days post intubation (top), and 30 day trends of dynamic compliance between survivors and non–survivors (bottom) indicating decreasing compliance over time with a steeper decline among non–survivors. Trends per compliance category are presented in supplement.

**Figure 4.**
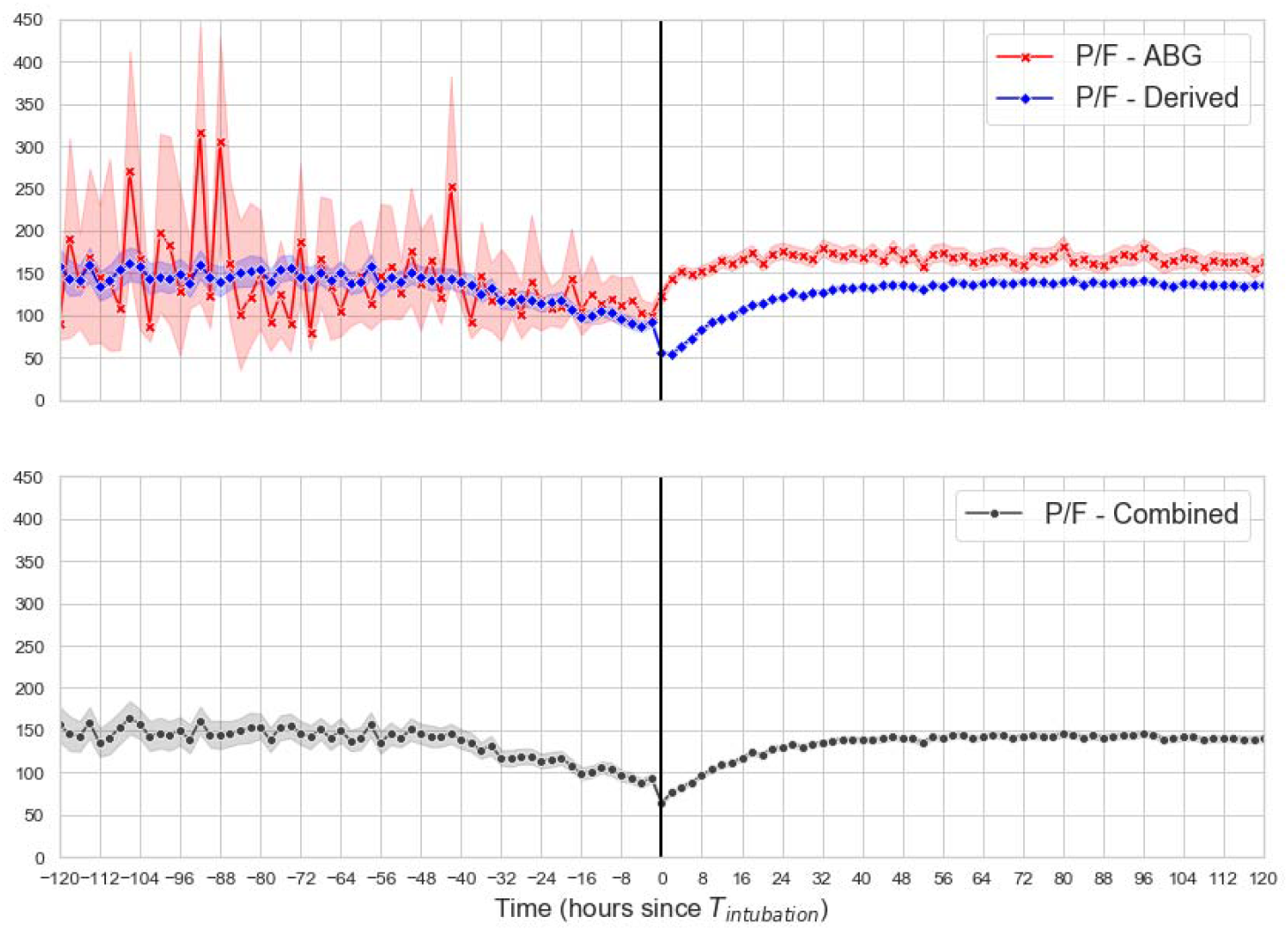
Trend over time in derived (from SpO2 from peripheral pulse oximetry) versus measured (from PaO2 in ABG) P/F ratio. The vertical black line denotes intubation time (Ti). Shaded areas indicate variability in measurements due to many missing measured PaO2 values relative to continually available SpO2 values. However, the direction of change over time is similar in derived and measured P/F values. The gap in derived and measured P/F during the first 24 hours of mechanical ventilation likely represents a combination of the maximum SpO2 being 100% (as opposed to PaO2 which can be over 600) which sets an upper limit to the derived P/F from SpO2; and due to the shape of the oxygen dissociation curve wherein small changes in SpO2 correspond to larger changes in PaO2.

**Figure 5.**
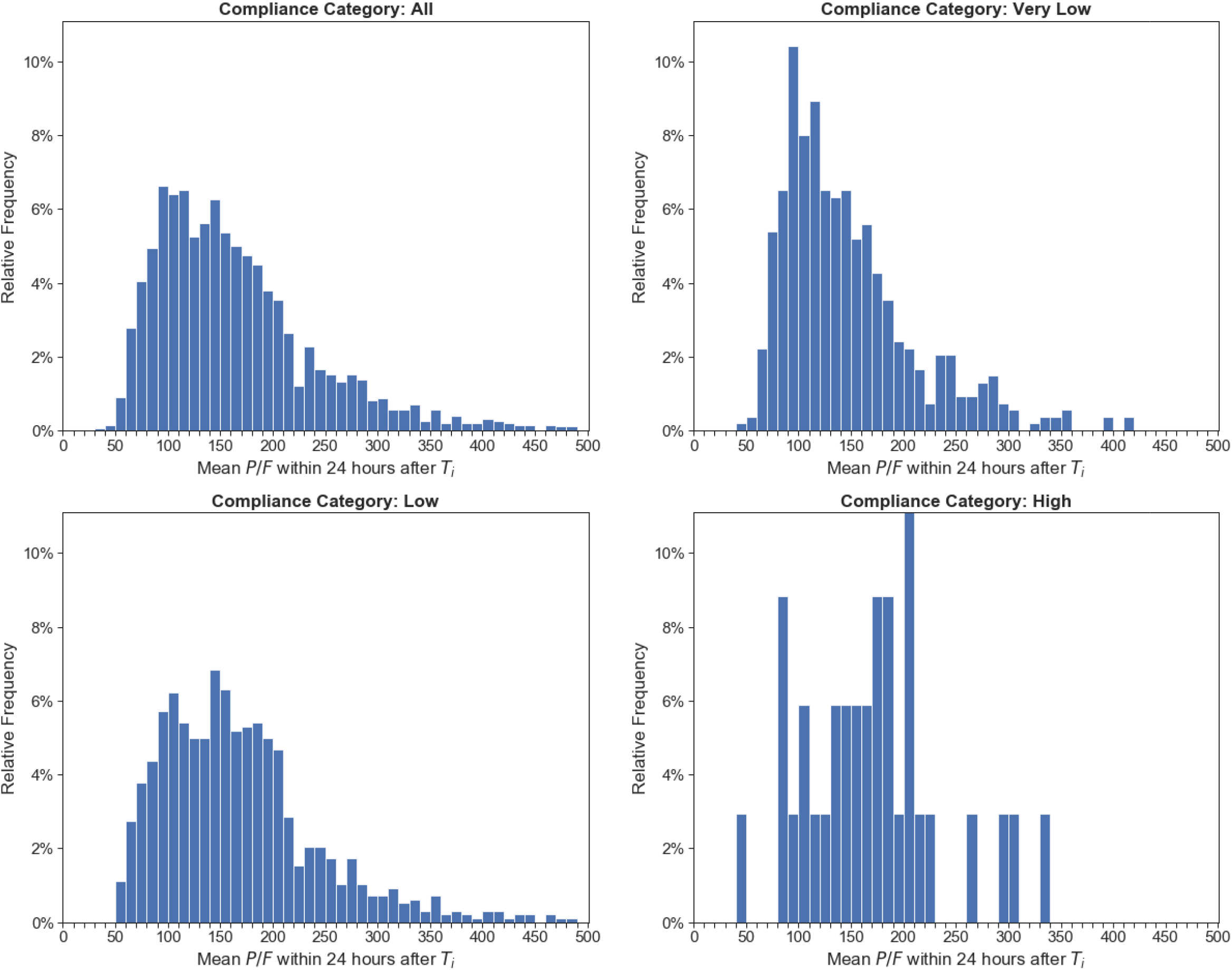
Frequency of ABG P/F by compliance category

In the 12 hours post intubation, the mean ABG P/F ratio was 155·33 (SD 88·47) for the overall group, and similar across groups. (Figure 2) Those in the very low compliance categories received higher FiO_2_ for longer periods of time prior to intubation (in the setting of also having longer average time to intubation). Prior to intubation, the group with normal to high compliance were exposed to FiO_2_ > 60% for 57·53 hours (IQR 6·68, 181·66) compared to 12·82 hours (IQR 0·45, 44·59) in the low-normal category (Table 3).

### Duration of intubation

The average duration of intubation was 14·25 days (SD13·69). Among those who survived, median duration was 11·9 days (IQR 4·8, 29·3) and mean was 18·26 (STD 16·91) days. Among those who died, median duration was 8·8 days (IQR 4·0, 17·1) and mean was 12·32 (STD 11·35). It should be noted that the length of intubation for survivors is an underestimation due to the fact that 19.8% of survivors were discharged while still mechanically ventilated.

The general trend of derived P/F ratios paralleled the ABG P/F ratios prior to intubation, although with high degree of variability among the ABG P/F ratios prior to intubation (wide 95% CI, shaded gray), due to many ABGs not being performed. Post-intubation, where many more ABGs were drawn, the two curves diverge for the first 48 hours, and then trend together over time. (Figure 4)

### Oxygenation Index (OI)

The mean OI for the entire cohort in the 24 hours after intubation was 11·12 (5·67), and was slightly worse in the very low compliance group 12·29 (5·70).

### Lung Mechanics and Ventilator Settings

Lung compliance for the whole cohort decreased over time, with a steeper trajectory among those who died (Figure 3). This was seen more clearly in the low-normal compliance group and high compliance groups likely secondary to the ‘floor effect’ (very low compliance numbers starting at a very low value) (Figure S·4). On average, patients received 6·9 cc/kg (SD 1·2) of ideal body weight as the ventilator setting. (Table 4) As expected, the very low lung compliance group had the highest average peak airway pressure, plateau pressure, and resulting driving pressures. The mean driving pressure for the whole cohort was 16·24 (SD 6·37), and 20·54 (SD·77) for the very low compliance group compared to 13·38 (SD 3·88) for the low-normal compliance group.

**Table 4.**
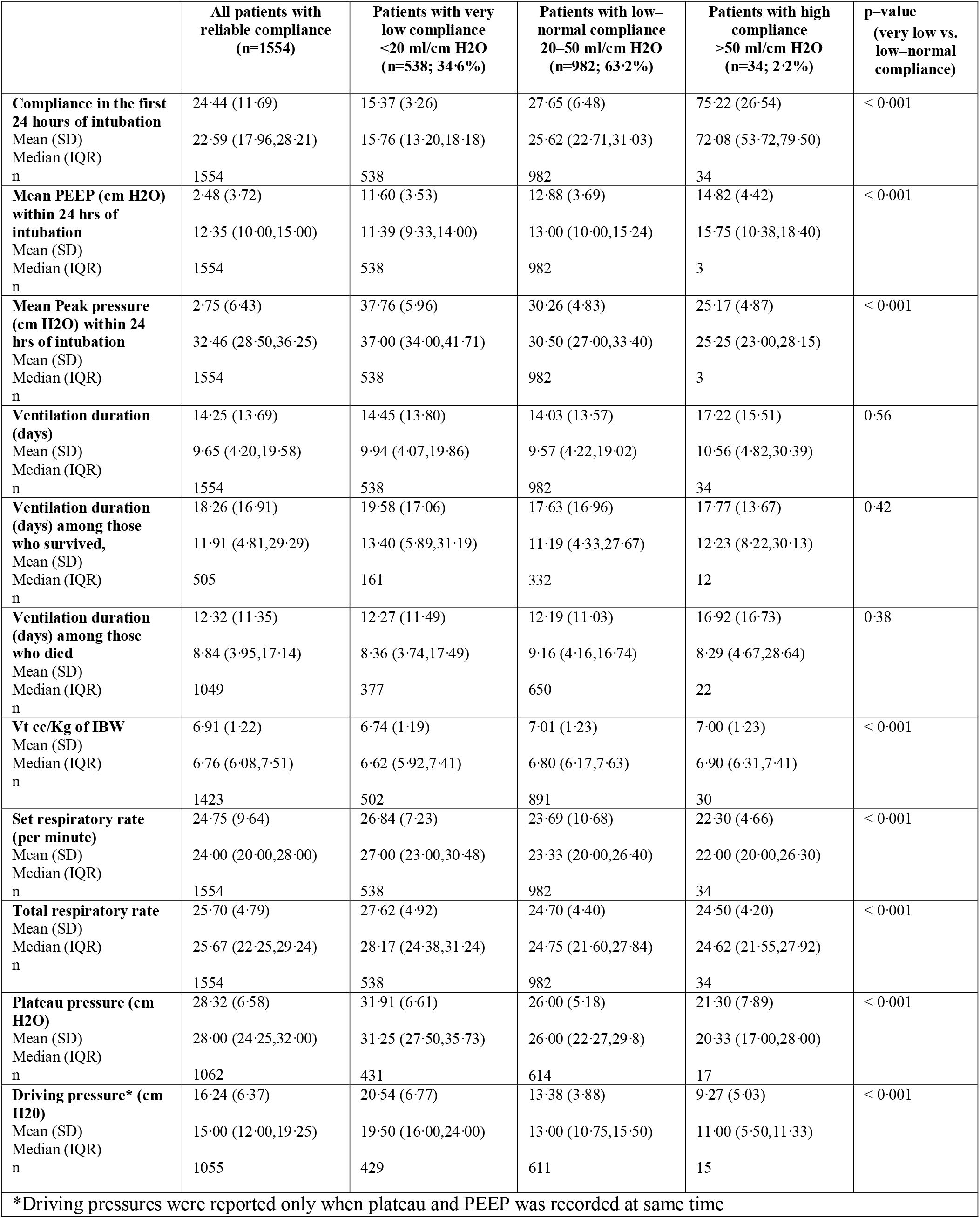
Mechanical ventilator obtained parameters (lung mechanics and ventilator settings)

### Proportion of Deaths and Discharge to Home

Table 5 presents the disposition status of patients based on the index hospitalization which was available for all patients (unknown for 2 patients). Overall, of the 1554 patients, 67·5% (n=1049) died during the index hospital stay. Of the 505 patients who survived to hospital discharge, 100 (19.8%) were discharged while still on a mechanical ventilator. Of those who survived, 44·4% (n=224) were discharged home and the rest to rehabilitation or longer term care facilities. The very low compliance group had the highest mortality (70·1% versus 66·2%) and fewer survivors were discharged home (41·0% versus 46·0%).

**Table 5.** Hospital Mortality and Discharge location stratified by compliance group

|  | **All patients with reliable compliance**<br>**N=1554** | **Patients with very low compliance**<br>**<20 ml/cm H<sub>2</sub>O**<br>**(n=538; 34·6%)** | **Patients with low-normal compliance**<br>**20–50 ml/cm H<sub>2</sub>O**<br>**(n=982; 63·2%)** | **Patients with high compliance**<br>**>50 ml/cm H<sub>2</sub>O**<br>**(n=34; 2·2%)** | **p-value**<br>**(very low vs. low-normal compliance)** |
| --- | --- | --- | --- | --- | --- |
| **Deceased** % (n) | 67·50% (1049) | 70·07% (377) | 66·19% (650) | 64·71% (22) | 0·122 |
| **Survived** % (n) | 32·50% (505) | 29·93% (161) | 33·81% (332) | 35·29% (12) | 0·051 |
| **Discharged while on mechanical ventilator**, % of the survivors (n) | 19·80% (100) | 22·98% (37) | 17·77% (59) | 33·33% (4) | 0·171 |
| **Discharged home**, % of the survivors (n) | 44·36% (224) | 40·99% (66) | 46·08% (153) | 41·67% (5) | 0·286 |
| **Discharged to another facility including acute care and longer-term rehabilitation**, % of the survivors (n) | 55·25% (279) | 59·01% (95) | 53·31% (177) | 58·33% (7) | 0·233 |

## Discussion

Patients with COVIDARDS in the NorthCARDS dataset had heterogeneous lung compliance, as measured in the first 24 hours of intubation. Three observations were particularly notable and include the longer time to intubation for patients with very low lung compliance, the steeper trajectory of compliance decrease seen among those who died, and the severity of hypoxemia in those with high lung compliance. As others have noted, the course of COVID19 pneumonia and ARDS appears to start with a highly compliant lung but with profound hypoxemia.^11,12^ Therefore, it is possible that ARDS patients with low compliance detected at the time of intubation may well have started with a normal lung compliance with deterioration during the course of illness, in part due to the disease process itself, and possibly due to treatments administered. For example, it is possible that prolonged exposure to high concentrations of oxygen contributed to the low compliance seen once patients were intubated. High concentrations of oxygen have been demonstrated to cause lethal lung injury in animal models,^13–15^ and have been associated with increased mortality,^16,17^ severe lung injury, and pneumonia^18^ in humans. A recent study linked hyperoxia to microbial dysbiosis in both the lung and gut microbiome which could contribute to the lung injury.^19^ It is unclear whether earlier intubation, and/or lower oxygen saturation thresholds would have mitigated worsening of lung compliance. The recent ICU-ROX study did not find that conservative oxygen thresholds (SpO_2_ 90-97%) decreased ventilator days in intubated ICU patients,^20^ and the recent LOCO2 trial, conservative therapy (SpO_2_ 88-92%) was associated with increased mortality among intubated patients.^21^ However, these results may not apply to non-intubated patients. Many COVID-19 patients who were maintained without intubation had uniquely preserved mentation despite very low SpO_2_ levels (likely due to right-shifted oxygen dissociation curves) and did not meet conventional thresholds for intubation. Alternatively, clinicians have posited that PSILI (patient self-induced lung injury)^22^ due to extreme respiratory drives could exacerbate lung damage in COVID-19 disease. Prior to intubation patients were not receiving sedation and strong respiratory drives may have contributed to the lower lung compliance seen due to PSILI. Of course, these patients could have had very low compliance at the time of hospital presentation. In addition, the persistence of active disease itself could have led to progressively lower compliance due to persistent severe inflammation. A prospective study that includes a surrogate measure for compliance prior to intubation, ideally with serial measurements over time, and documentation of progression of ventilation and perfusion mismatch (including ultrasound or other radiography and dead space estimation) will help answer these questions. These investigations are relevant for ARDS in general and learnings will have implications for the management of ARDS beyond COVID19.

Degree of V/Q mismatch and hypoxemia does not appear to correlate with lung compliance, which corresponds to what colleagues have found in the non-ARDS analyses.^23^ Indeed, 42% of the cohort with high lung compliance in non-COVID ARDS patients had P/F levels under 150, which is similar to our findings in COVIDARDS. The extremely low P/F ratio, P/FP and high Oxygenation Index seen among patients in the high compliance group suggests ventilation perfusion mismatch which could be explained by the extensive micro-thrombi that have been reported, and the involvement of the vascular endothelium with impaired hypoxic pulmonary vasoconstriction.^24^ Questions have been raised about whether COVIDARDS should be treated differently than non-COVIDARDS. The more relevant question seems to be whether ARDS management should be different for patients with different severity of lung compliance impairment and different degrees of ventilation and perfusion mismatch. An index that takes into account oxygen impairment and compliance over time, pointing to predominance of dead space ventilation (thrombi) versus shunt physiology (alveolar and parenchymal pathology, and impaired vascular hypoxic vasoconstriction) may help clinicians tailor treatments for individual patients with ARDS.

Only 2·2% of patients were in the high compliance category (low elastance/ phenotype “L”).^25^ This is lower than the 12% reported in the recent secondary analysis of the LungSAFE data of non-COVID ARDS patients.^23^ However, it is important to note that our description of compliance variability is limited to ARDS patients who are mechanically ventilated. Many patients who met ARDS criteria based on hypoxemia and bilaterality of infiltrates did not receive mechanical ventilation until several days after admission. This period was likely prolonged compared to other viral pneumonia causes of ARDS due to the relatively preserved mental status in COVID-19 patients despite profound hypoxemia. Comparisons between studies need to consider the timing of intubation relative to symptom onset, and different practice patterns regarding thresholds for intubation. Disparate outcomes reported internationally are likely explained in large part by different comorbidity burden, severity of hypoxemia on hospital presentation, and different practice patterns regarding timing of intubation.

The strengths of this study include being the largest sample of COVIDARDS patients in a single health system which has granular patient-level data regarding respiratory mechanics and oxygenation. We have described methods for leveraging real-world data to determine lung compliance data in the absence of patient effort which could either over- or under-estimate true pressures. Our large sample size allowed us to maintain 1554 patients in the dataset who had reliable data on pulmonary mechanics.

Limitations of the present study are inherent to the retrospective nature of this data extraction from the electronic health record. We are unable to ensure that there was no significant airway resistance contributing to the measurement of dynamic compliance, and to account for the contribution of abdominal pressures and chest wall stiffness. The small and consistent margin of difference between static and dynamic compliance seen suggests that airway resistance contributed minimally to measured dynamic airway pressures. We assumed that the difference between dynamic and static compliance would be < 10 mL/ cm H_2_O due to airway resistance not being commonly observed in the early stages of COVIDARDS. In non-COVID-19 related ARDS the mean difference between peak and plateau pressures has been found to be 6-7 cmH_2_O.^26^ However, given that 50% (n=804) of patients had a BMI of over 30, it is possible that chest wall compliance contributed to a decreased measured compliance in some patients. A further limitation is our inability to control for factors which influenced decisions about timing of intubation for COVID19 patients. For example, those who were intubated earlier may have had altered mental status which could confound differences seen in mortality associated with lung compliance. Limits to resuscitation due to patient and family preference have also not been presented in this descriptive analysis. These factors will need to be accounted for in future inferential studies

In summary, we present the methods for establishing the NorthCARDS dataset of COVIDARDS patients, and the range of lung compliance and oxygen trajectories seen in these patients. These data will inform phenogrouping research to further understand COVIDARDS towards tailored approaches to treatment which maybe also be applicable to non-COVID-19 related ARDS.

## Data Availability

The authors welcome qualified third parties and experts to review the paper and would gladly share the de-identified dataset as well research methods and data analysis.

## Authors’ Contributions

Daniel Jafari - literature search, figures, study design, data collection, data interpretation, writing

Amir Gandomi - figures, study design, data collection, data analysis, data interpretation, writing

Alex Makhnevich - literature search, figures, study design, data collection, data interpretation, writing

Michael Qiu - data collection, data analysis, data interpretation

Daniel Rolston - study design, data collection, data interpretation, writing

Eric Gottesman - study design, data collection, data interpretation, writing

Adey Tsegaye - study design, data collection, data interpretation

Paul Mayo - study design, data interpretation

Molly Stewart - literature search, data collection, writing

Meng Zhang - study design, data collection, data interpretation

Negin Hajizadeh - literature search, figures, study design, data collection, data interpretation, writing

All authors have had access to and verified the underlying data. In Particular DJ, NH, AM, AT, EG independently chart reviewed random selections of patients to verify accuracy of data.

## Data sharing statement

|  |  |
| --- | --- |
| Will individual participant data be available (including data dictionaries)? | Yes |
| What data in particular will be shared? | Individual participant data that underlie the results reported in this article, after de-identification (text, tables, figures, and appendices) |
| What other documents will be available? | Study protocol, statistical analysis plan, analytic code |
| When will data be available (start and end dates)? | Beginning 3 months and ending 36 months following article publication |
| With whom? | Investigators whose proposed use of the data has been approved by an independent review committee (Northwell COVID Consortium) identified for this purpose |
| For what types of analyses? | For individual participant data meta-analysis, or to achieve the aims in the approved proposal |
| By what mechanism will data be made available? | Proposals may be submitted up to 36 months following article publication<br>After 36 months the data will be available in our university's data warehouse but without investigator support other than deposited metadata<br>Information regarding submitting proposals and accessing data may be found at (link to be provided) |

## Acknowledgments

We wish to acknowledge all of the staff at the hospitals providing compassionate care to patients in the midst of this pandemic. This includes the respiratory therapists (RTs), registered nurses (RNs), doctors (MDs), all of the support staff, and the patients and their families who suffered durig this particularly difficult period, and to the Northwell ARDS Collaborative and the COVID19 Research Consortium who worked diligently to better understand this disease from the data collected, in order to inform each other of risk factors for poor outcomes and ideas for how we can work to improve outcomes.

## Supplement

### Data Assumptions and Structuring

#### Height and Weight

Patient’s recorded height and weight during the hospital visit was used. If this data was missing we obtained data from prior inpatient or outpatient records with the assumption that over a 2 year period height and weight remained stable.

#### Time of intubation (T_i_)

To accurately capture the time of intubation (Ti) the earliest of the following data points were used: an endotracheal intubation procedure note in the electronic medical record; “Start Time” of mechanical ventilation documented in the mechanical ventilation record; time of administration of rapid sequence induction medications (cisatracurium, rocuronium, vecuronium, succinylcholine, etomidate); documentation of ventilator settings by the respiratory therapist in the mechanical ventilation record; nursing documentation of ventilator use in the oxygen delivery method section of the flowsheets; or initiation of continuous infusions of sedative medications (fentanyl, propofol).

#### Duration of intubation

The duration of intubation was calculated by identifying the extubation time with the following criteria in the electronic medical record using a step-wise approach. First we searched for the field “time ventilator discontinued” in the mechanical ventilation record. If that data point was unavailable we used the “reason ventilator discontinued” or “ventilator mode off” fields, followed by time of “airway removal note”. In patients where there was evidence of mechanical ventilator records being discontinued, but the above evidence of extubation was missing, the first instance of supplemental oxygen delivery by methods not possible via mechanical ventilation was used. If the patient expired while receiving mechanical ventilation, expiration time was used as time of extubation within 8 hours of last mechanical ventilator record. Finally, if none of the aforementioned exhaustive criteria were found, we added 4 hours to the last set of ventilator parameters recorded in the mechanical vent record and assumed that as the time of extubation. The duration of invasive mechanical ventilation was defined as the number of days between T_i_ and successful extubation without the need for reintubation within 48 hours, or death.

#### ICU readmission

Intensive care unit (ICU) readmission was defined as over 48 hours between ICU discharge and ICU readmission and the first visit was used for this analysis. During the surge, hospitals within the system have expanded critical care capacity by creating dedicated COVID ICUs at certain hospitals.

#### Oxygen Delivery Method, Concentration, and Degree of Hypoxemia

The FiO_2_ delivered was calculated based on the following formula: for nasal cannula or non-rebreather face mask, each liter of oxygen flow added 0·04 to 0·21 (room air), with a maximum of 6 liters per minute for nasal cannula and 15 liters per minute for non-rebreather mask. In cases where both nasal cannula and non-breather mask was used at the same time, FiO_2_ was assumed to be 1·0. For non-invasive ventilation (NIV), high flow nasal cannula (HFNC), or venturi mask, the documented FiO_2_ was used. Of note, there was limited use of NIV or HFNC because of concerns for airborne transmission of COVID-19 during this particular time. In the instances where the delivery method was not recorded in the electronic medical record, the previous recorded method was presumed to be have been continued, until change in flow rate or delivery method was noted.

To be able to accurately map hypoxemia prior to intubation, we used both arterial blood gas data on partial pressure of oxygen (PaO_2_) and peripherally measured oxygen saturation (SpO_2_). We calculated SpO_2_:FiO_2_ ratios as well as PaO_2_:FiO2 ratios over time for each patient across their entire hospital stay. For separate analyses we converted SpO_2_:FiO_2_ to PaO_2_:FiO_2_ ratios (‘derived P/F’) to obtain an estimated trajectory of PaO_2_ over time (refs). The assumption that derived P/F would have parallel trends compared to ABG derived P/F was tested.

#### Respiratory System Compliance

We used both static compliance (change in lung volume per unit change in pressure in the *absence of flow)* using the plateau pressure recorded in the electronic medical record, (Tidal Volume/ [Plateau Pressure – Peak End Expiratory Pressure (PEEP]); and dynamic compliance using the Peak Inspiratory Pressure (PIP) (change in lung volume per unit change in pressure in the *presence of flow)*, (Tidal Volume/ [PIP – PEEP]) when patients were deeply sedated/paralyzed as described below. We only included values obtained at the time of full patient sedation, which was defined as the administration of intermittent bolus or continuous infusions of paralytics (cisatracurium, rocuronium, vecuronium) and (patient respiratory rate - set respiratory rate) <2 (Figure S·7). We made the assumption that patients would not have a significant component of airway resistance for most COVID-19 respiratory failure patients in the early stage of disease (no more than a difference of 5-7 mmHg between PIP and Plateau pressures), and that therefore this added pressure due to flow would have a minimal contribution to overall measured compliance. This assumption was tested by visualizing the difference between static and dynamic compliance seen over time (Figure S·3).

#### Interventions to prevent the worsening of ARDS

We recorded the use of paralytic medications, vasopressor or inotropic medications, lung protective ventilatory settings, steroids, and various experimental treatments that were provided during the surge (azithromycin, vitamin C, hydroxychloroquine, tocilizumab, anakinra, steroids) (Table 2). Proning was defined as a binary variable, and was extracted by string pattern search of items related to the patient position in the EMR. The search discerns positive matches (e,g., “proned”) from the negative ones (e.g., “unproned”). We created a standardized set and returned tidal volumes for each patient based on predicted body weight, calculated based on sex and height.

**Supplementary Table S1.** Demographics for the group of patients with no reliable compliance measurements in the first 24 hours compared to 1979 COVID–ARDS database

|  | **All patients**<br>**N=1979** | **No reliable compliance in first**<br>**24hrs.**<br>**N= 425** |
| --- | --- | --- |
| **Age, median (IQ)** | 66·0 [57·0,74·0] | 69·0 [61·0,77·0] |
| **Female n (%)** | 633 (32·0) | 143 (33·6%) |
| **Race** |  |  |
| **Black/African American** | 364 (18·4) | 74 (17·4) |
| **Asian** | 189 (9·6) | 35 (8·2) |
| **Native Am/Alaskan** | 4 (0·2) | 2 (0·5) |
| **Native Hawaii/Pacif Isl** | 3 (0·2) | 1 (0·2) |
| **Other/Multiracial** | 589 (29·8) | 109 (25·6) |
| **Unknown** | 106 (5·4) | 25 (5·9) |
| **White** | 724 (36·6) | 179 (42·1) |
| **Comorbidities (Charlson Index)**<br>**mean (SD)** | 5·1 (3·3) | 6 (3·4) |
| **MEWS on admission mean (SD)** | 4·1 (1·9) | 4·2 (1·9) |
| **Chronic Lung Disease n (%)** | 140 (7·1) | 38 (8·9) |
| **Diabetes n (%)** | 887(44·8) | 212 (49·9) |
| **HTN n (%)** | 1292 (65·3) | 280 (65·9) |
| **CHF n (%)** | 162 (8·2) | 48 (11·3) |
| **CAD (%)** | 280 (14·1) | 72 (16·9) |
| **CKD (%)** | 302 (15·3) | 74 (17·4) |
| **ESRD n (%)** | 113 (5·7) | 28(6·6) |
| **Positive Smoking n (%)** | 240 (15·4) | 45 (14·8) |
| **Malignancy n (%)** | 199 (10·1) | 52 (12·2) |
| **BMI on admission mean (SD)** | 31·5 (34·3) | 33·0 (73·2) |
| **BMI Categories n (%)** |  |  |
| **BMI <18 underweight** | 23 (1·2) | 13 (3·1) |
| **BMI 18 – <30 normal–overweight** | 964 (48·7) | 224 (52·7) |
| **BMI 30 – <40 obese** | 676 (34·2) | 123 (28·9) |
| **BMI >=40 extremely obese** | 316 (16) | 65 (15·3) |
| **Median hours from hospital**<br>**presentation to intubation (IQR)** | 47·0 (5·9,121·4) | 31·9 (1·7,107·1) |
| **Count of patients intubated within**<br>**48 hours of presentation (%)** | 995 (50·28) | 243 (57·2) |
| **Pre–intubation O2 supplementation n (%)** |  |  |
| **NRB** | 995 (50·3) | 278 (65·4) |
| **NRB+NC** | 47 (2·4) | 6 (1·4) |
| **NC** | 125 (6·3) | 43 (10·1) |
| **HFNC** | 43 (2·2) | 10 (2·4) |
| **NIV (BiPAP+CPAP)** | 60 (3) | 9 (2·1) |
| **Venturi** | 19 (1) | 4 (0·) |
| **Other** | 211 (10·7) | 75 (17·6) |

**Supplementary Table S2.** Oxygenation trends and duration of ventilation for group with no reliable compliance data, compared to total cohort.

|  | **Total Cohort**<br>**N= 1979** | **All patients with reliable**<br>**compliance**<br>**N=1554** | **No reliable compliance in first**<br>**24hrs**<br>**N=425** |
| --- | --- | --- | --- |
| **P/F derived <u>pre</u> intubation (12hr mean)**<br>Mean (SD)<br>Median (IQR)<br>n | 98.34 (89.22)<br>60.49 (53.15,107.02)<br>1772 | 94.64 (85.08)<br>60.13 (52.17,98.92)<br>1419 | 113.20 (103.02)<br>63.43 (56.08,138.10)<br>353 |
| **P/F derived <u>post</u> intubation, (12hr mean)**<br>Mean (SD)<br>Median (IQR)<br>n | 68.96 (41.48)<br>53.81 (40.65,83.85)<br>1954 | 68.20 (38.77)<br>54.32 (40.97,83.07)<br>1543 | 71.79 (50.31)<br>51.64 (40.04,86.71)<br>411 |
| **P/F from <u>ABG</u> <u>pre</u> intubation (12hr mean)**<br>Mean (SD)<br>Median (IQR)<br>n | 113.63 (113.62)<br>76.23 (61.73,113.33)<br>556 | 109.32 (100.27)<br>75.31 (61.56,110.72)<br>454 | 132.83 (159.33)<br>83.54 (63.55,127.53)<br>102 |
| **First ABG P/F post intubation (within 4 hours after Ti),**<br>Mean (SD)<br>Median (IQR)<br>n | 155.07 (127.70)<br>121.00 (86.00,188.92)<br>1376 | 149.39 (105.58)<br>120.00 (85.00,183.81)<br>1116 | 179.47 (194.54)<br>129.50 (90.00,212.59)<br>260 |
| **P/F from <u>ABG</u> <u>post</u> intubation (12 hr post, ABG)**<br>Mean (SD)<br>Median (IQR)<br>n | 160.78 (95.58)<br>138.33 (97.11,198.00)<br>1844 | 155.33 (88.47)<br>136.31 (96.50,189.79)<br>1484 | 183.21 (118.08)<br>150.17 (101.75,237.12)<br>360 |
| **P/F gradient <u>derived</u> 24 hrs <u>pre</u> intubation (derived P/F)**<br>Mean (SD)<br>Median (IQR)<br>n | 85.44 (1869.17)<br>-6.22 (-106.55,1.94)<br>1649 | -321.66 (2054.35)<br>-6.39 (-108.70,1.58)<br>1345 | -125.17 (500.91)<br>-4.82 (-101.83,3.96)<br>304 |
| **Proportion of time on FiO2>=60% <u>pre</u>-intubation**<br>Mean (SD)<br>Median (IQR)<br>n | 60.45% (37.27%)<br>70.40% (25.98%,97.86%)<br>1703 | 61.86% (36.79%)<br>73.31% (28.19%,97.91%)<br>1380 | 54.42% (38.76%)<br>60.11% (14.67%,96.88%)<br>323 |
| **Proportion of time on FiO2>=60% <u>post</u>-intubation**<br>Mean (SD)<br>Median (IQR)<br>n | 54.05% (35.13%)<br>51.45% (20.01%,93.69%)<br>1959 | 52.62% (34.62%)<br>48.78% (19.43%,89.65%)<br>1554 | 59.53% (36.55%)<br>60.45% (23.43%,100.00%)<br>405 |
| **PEEP (Mean within 24 hrs of Ti)**<br>Mean (SD)<br>Median (IQR)<br>n | 12.29 (3.87)<br>12.00 (10.00,15.00)<br>1957 | 12.48 (3.72)<br>12.35 (10.00,15.00)<br>1554 | 11.57 (4.32)<br>11.00 (8.58,15.00)<br>403 |
| **Mean Peak pressure (cm H2O) within 24 hrs of intubation**<br>Mean (SD)<br>Median (IQR)<br>n | 32.35 (6.85)<br>32.00 (27.71,36.20)<br>1953 | 32.75 (6.43)<br>32.46 (28.50,36.25)<br>1554 | 30.82 (8.10)<br>30.00 (25.50,35.00)<br>399 |
| **Vt cc/Kg of IBW**<br>Mean (SD)<br>Median (IQR)<br>n | 6.96 (1.28)<br>6.79 (6.10,7.60)<br>1791 | 6.91 (1.22)<br>6.76 (6.08,7.51)<br>1423 | 7.11 (1.46)<br>6.99 (6.23,7.84)<br>368 |
| **Set respiratory rate (per minute)**<br>Mean (SD)<br>Median (IQR)<br>n | 24.46 (13.92)<br>24.00 (20.00,27.60)<br>1961 | 24.75 (9.64)<br>24.00 (20.00,28.00)<br>1554 | 23.34 (24.03)<br>20.00 (17.60,25.17)<br>407 |
| **Total respiratory rate**<br>Mean (SD)<br>Median (IQR)<br>n | 25.59 (4.86)<br>25.54 (22.00,29.00)<br>1962 | 25.70 (4.79)<br>25.67 (22.25,29.24)<br>1554 | 25.17 (5.14)<br>25.25 (21.62,28.67)<br>40 |
| **Plateau pressure**<br>Mean (SD)<br>Median (IQR)<br>n | 27.91 (6.76)<br>27.50 (23.89,32.00)<br>1282 | 28.32 (6.58)<br>28.00 (24.25,32.00)<br>1062 | 25.91 (7.26)<br>25.00 (21.50,29.81)<br>220 |
| **Driving pressure**<br>Mean (SD)<br>Median (IQR)<br>n | 16.07 (6.35)<br>15.00 (12.00,19.00)<br>1266 | 16.24 (6.37)<br>15.00 (12.00,19.25)<br>1055 | 15.23 (6.19)<br>14.00 (11.71,17.83)<br>211 |
| **Oxygenation Index**<br>Mean (SD)<br>Median (IQR)<br>n | 11.05 (6.16)<br>9.95 (6.76,13.83)<br>1627 | 11.12 (5.67)<br>10.17 (7.00,13.90)<br>1341 | 10.70 (8.06)<br>8.86 (5.90,13.40)<br>286 |
| **PFP Value**<br>Mean (SD)<br>Median (IQR)<br>n | 101.28 (95.07)<br>76.69 (49.42,117.28)<br>1947 | 96.41 (73.98)<br>76.57 (50.86,113.73)<br>1554 | 120.51 (150.73)<br>77.73 (44.13,135.09)<br>393 |

|  | **Total N=1979** | **No reliable compliance n=425** |
| --- | --- | --- |
| **Deceased** % (n) | 69.48% (1375) | 76.71% (326) |
| **Survived** % (n) | 30.52% (604) | 23.29% (99) |
| **Discharged while on mechanical ventilator**, % of the survivors (n) | 20.20% (122) | 22.22% (22) |
| **Discharged home**, % of the survivors (n) | 43.54% (263) | 39.39% (39) |
| **Discharged to another facility including acute care and longer-term rehabilitation**, % of the survivors (n) | 56.13% (339) | 60.61% (60) |

**Figure S1.**
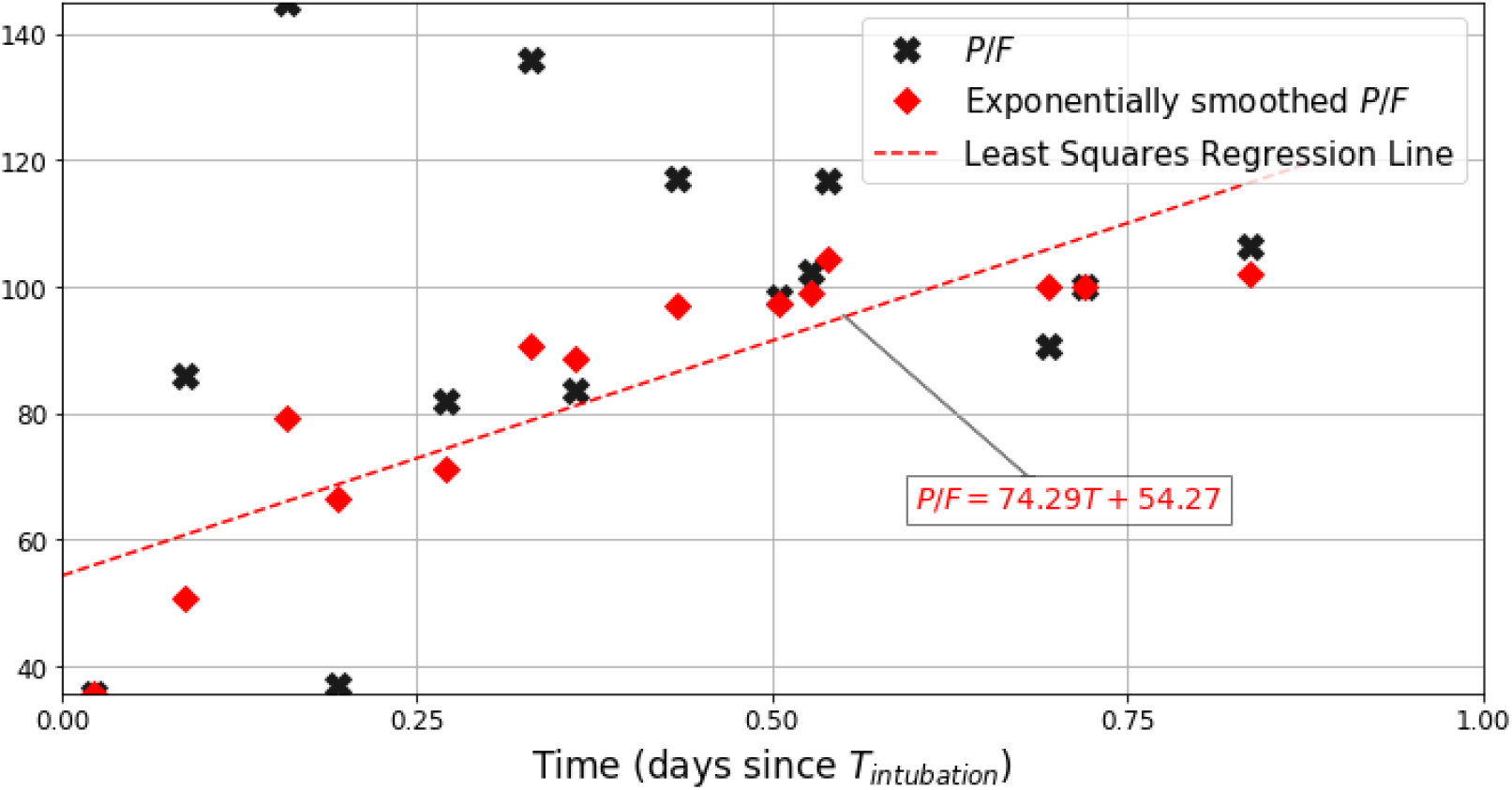
Trend calculation method for P/F ratio. First, we use exponential smoothing to reduce the noise in data (red diamond points). We then find OLS regression line passing through the smoothed points. Trend is the slope of the line (i.e., 74·29).

**Figure S2.**
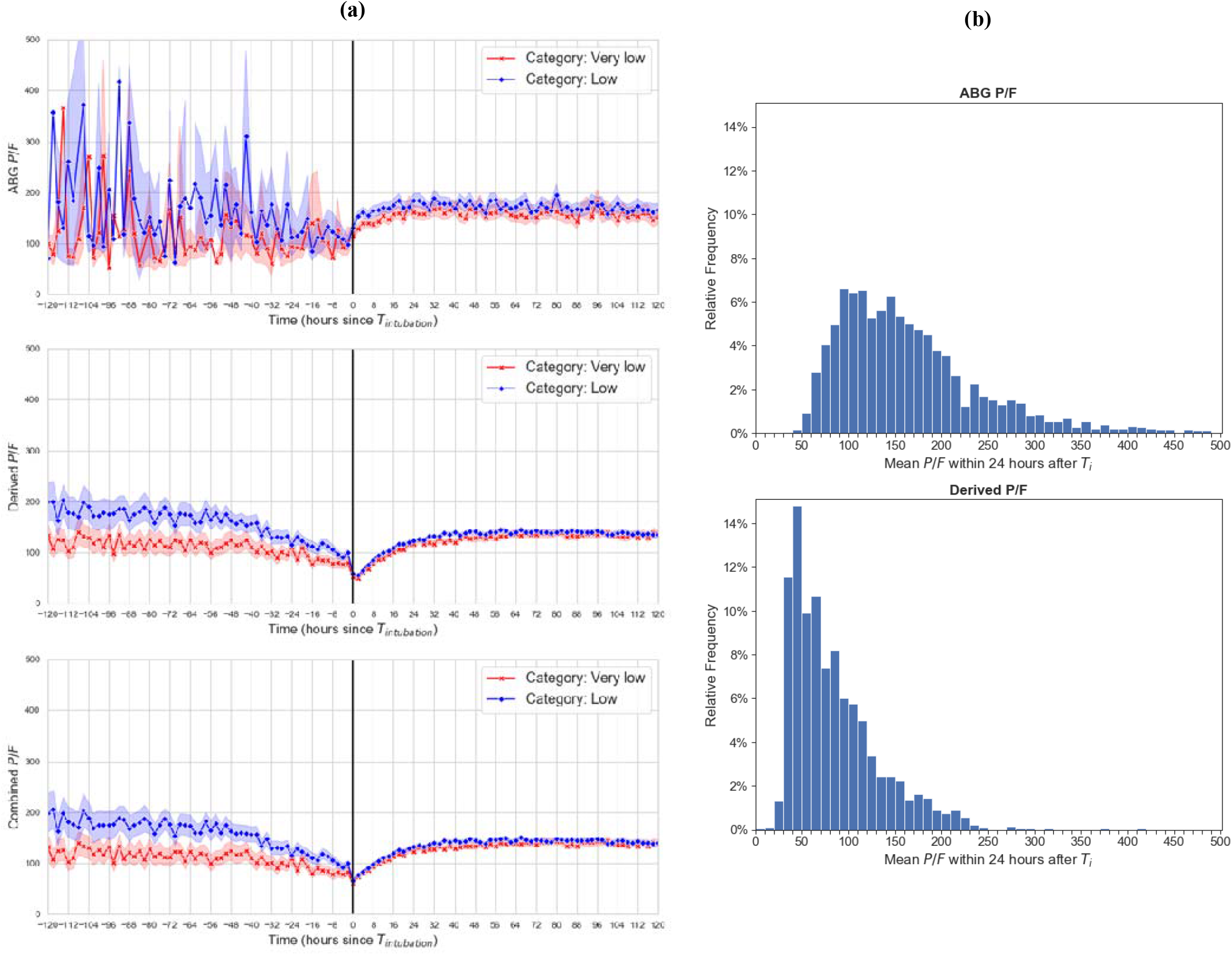
Comparison of derived P/F and ABG based P/F ratios. **(a)** Trends appear similar over time in very low and low-normal categories of lung compliance, and with ABG P/F (top), derived P/F from SpO2 (middle) and combining both methods with preference for ABG P/F (bottom). The ABG derived P/F has large variation due to fewer datapoints. The very low compliance category consistently has lower P/F ratio which is more pronounced pre–intubation. **(b)** P/F ratios in the first 24 hours of intubation are higher with ABG P/F (top) compared to derived ABG (bottom). Derived P/F underestimates the ratio (due to lower derived PaO2) due to oxygen dissociation curve where there is a greater range of PaO2 for a given SpO2, and the fixed upper limit of SpO2 at 100%.

**Figure S3.**
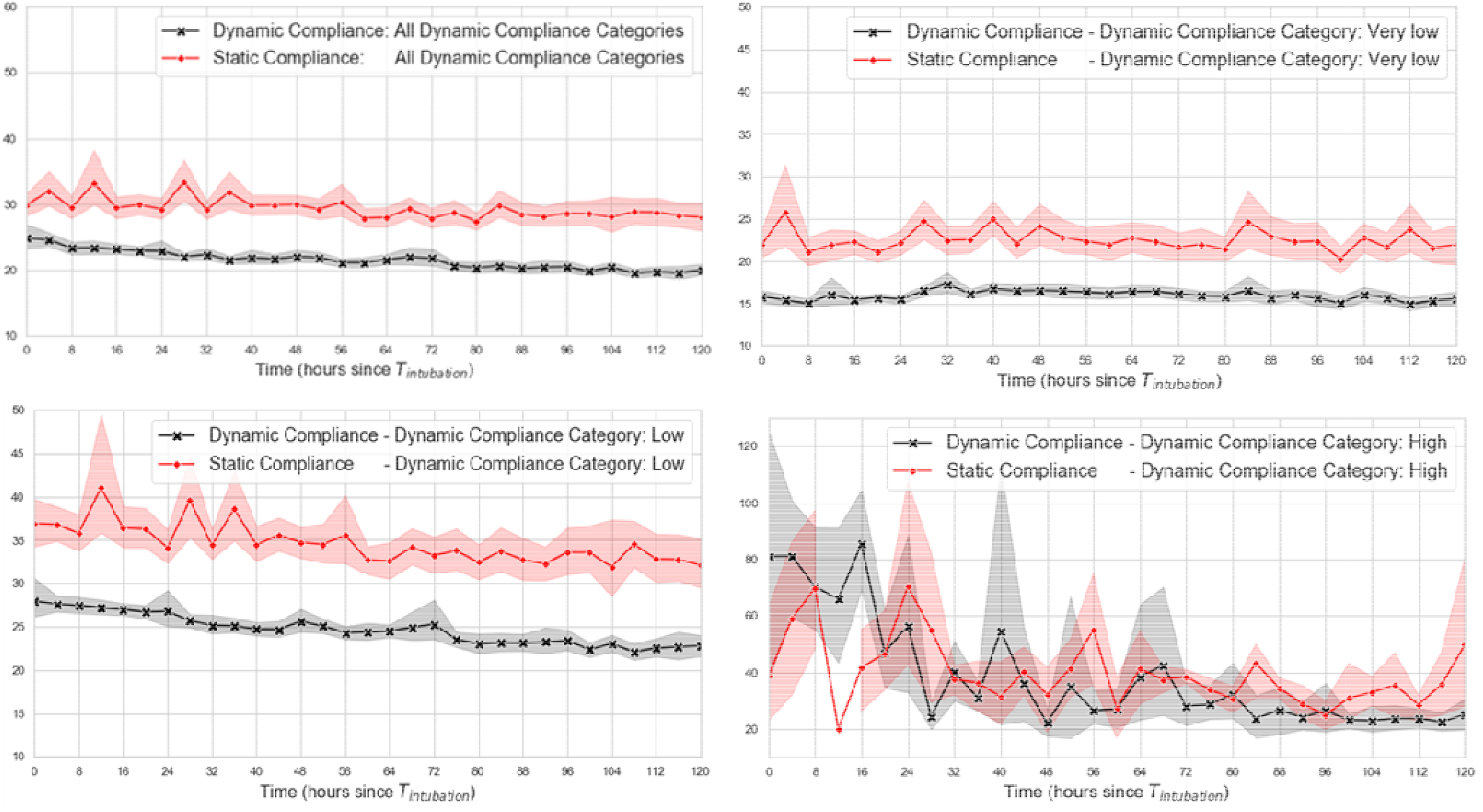
Static versus Dynamic Compliance over time to test the assumption that the dynamic compliance would be continuously lower than static with a relatively fixed difference over time, and regardless of compliance category.

**Figure S4.**
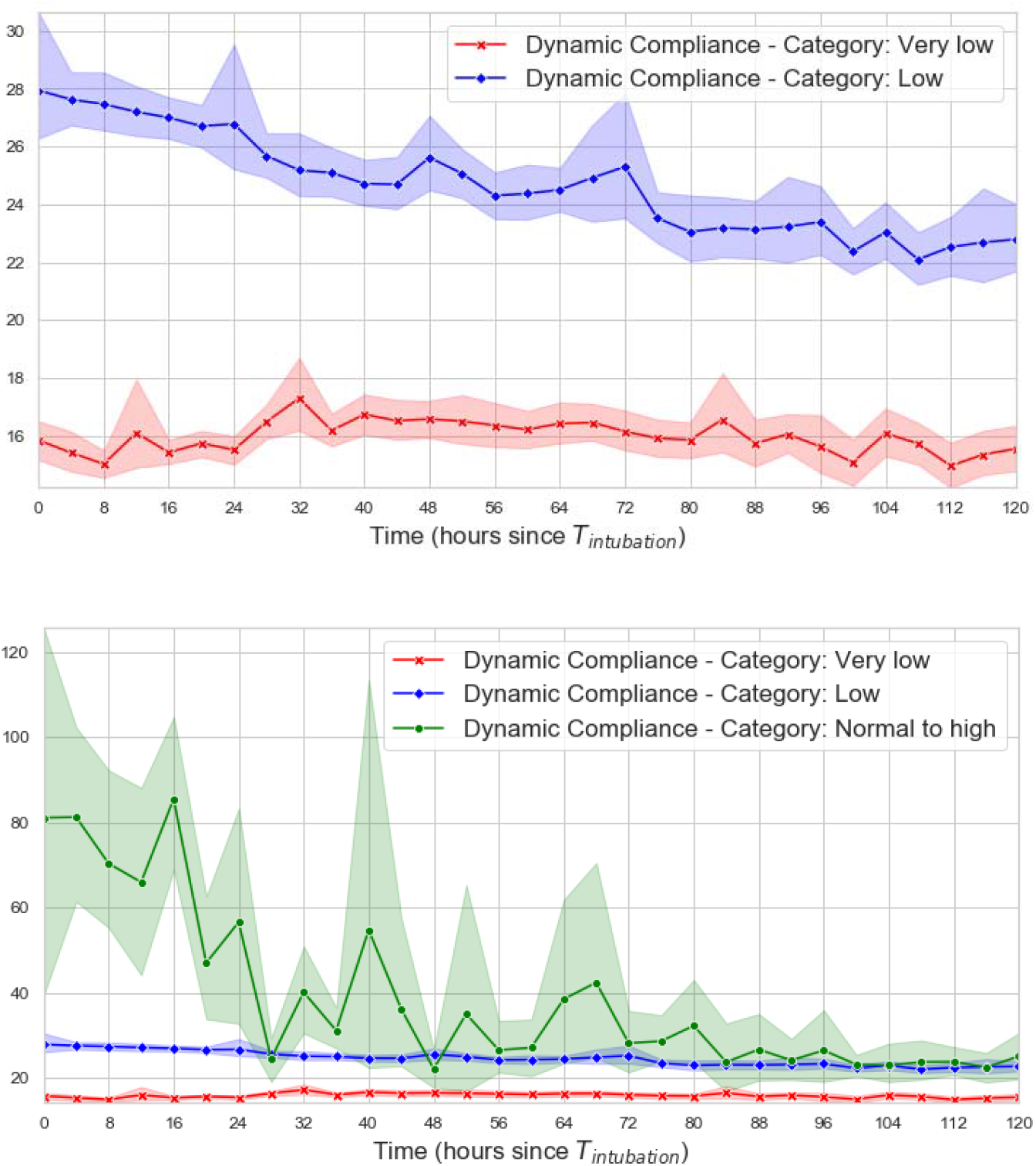
Comparison of Dynamic Compliance change over time (shaded areas represent 95% Confidence Intervals). A steeper trajectory is seen for the high and low–normal categories over time. Large variability seen in the high compliance group secondary to small sample size.

**Figure S5.**
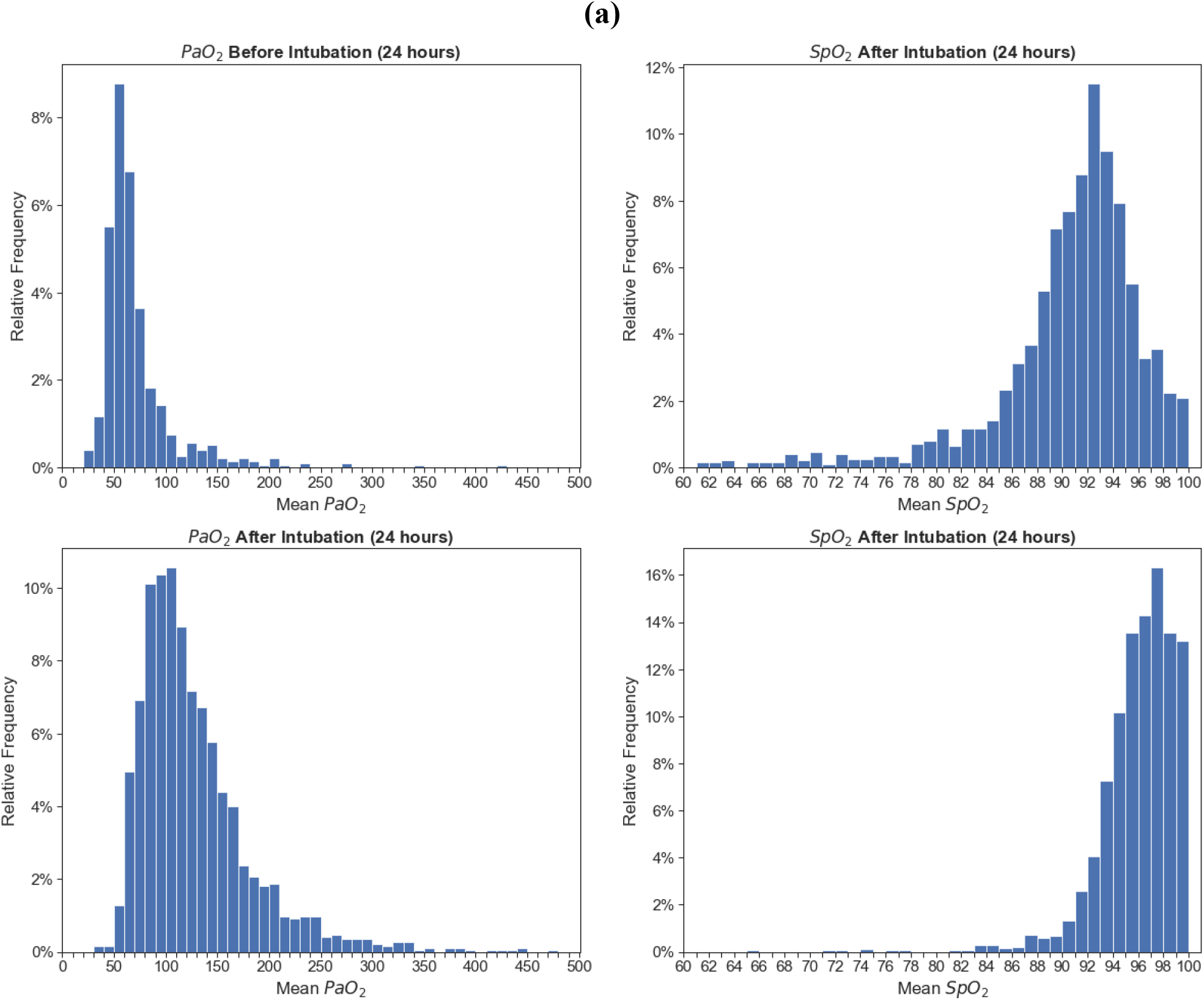
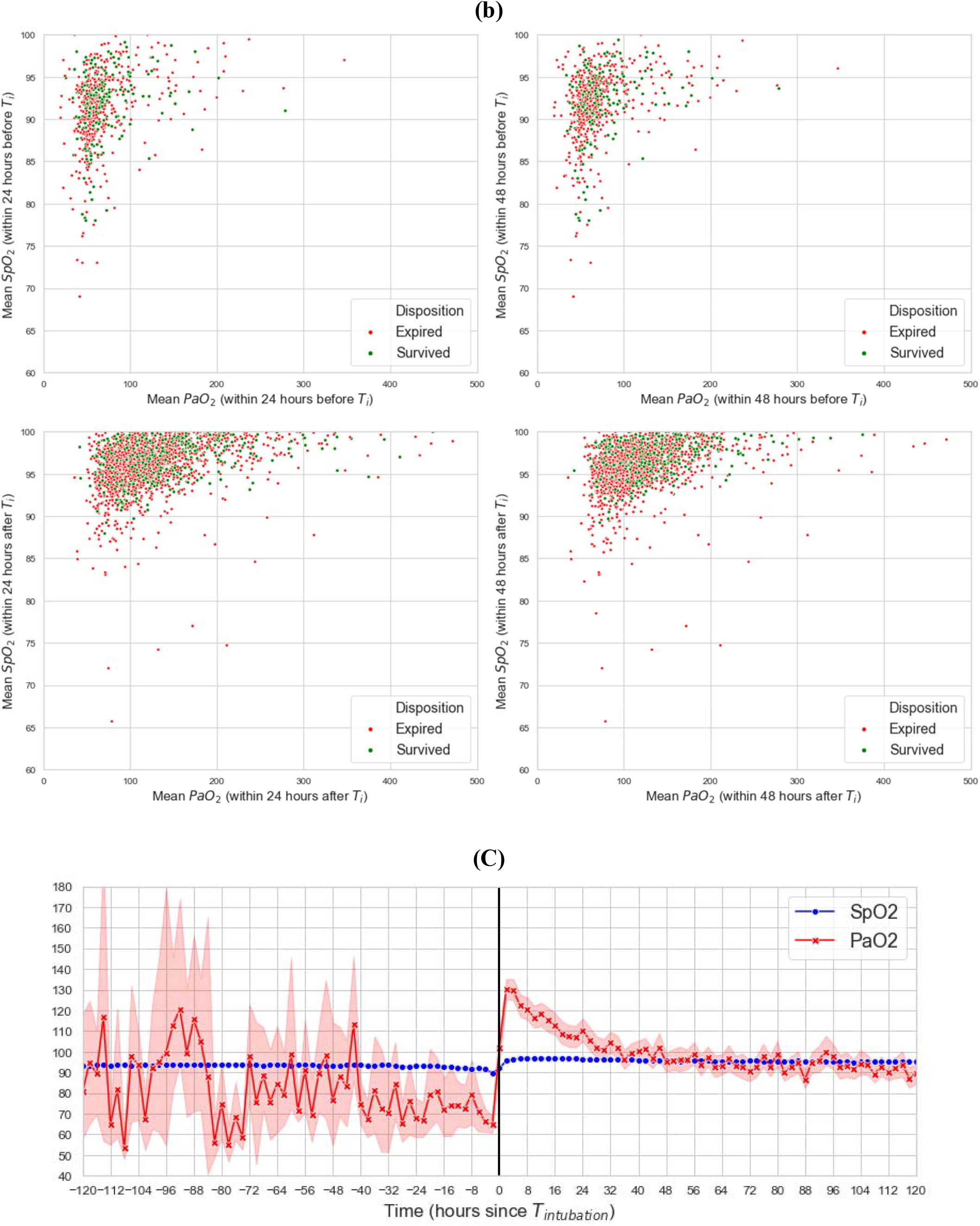
Distribution of PaO2 and SpO2 **a**. 24 hours before intubation (on left hand side) and after (on the right hand side) intubation (Ti) **b**. mean SpO2 and PaO2 association **c**. trend in SpO2 and PaO2 over the entire hospital course.

**Figure S7.**
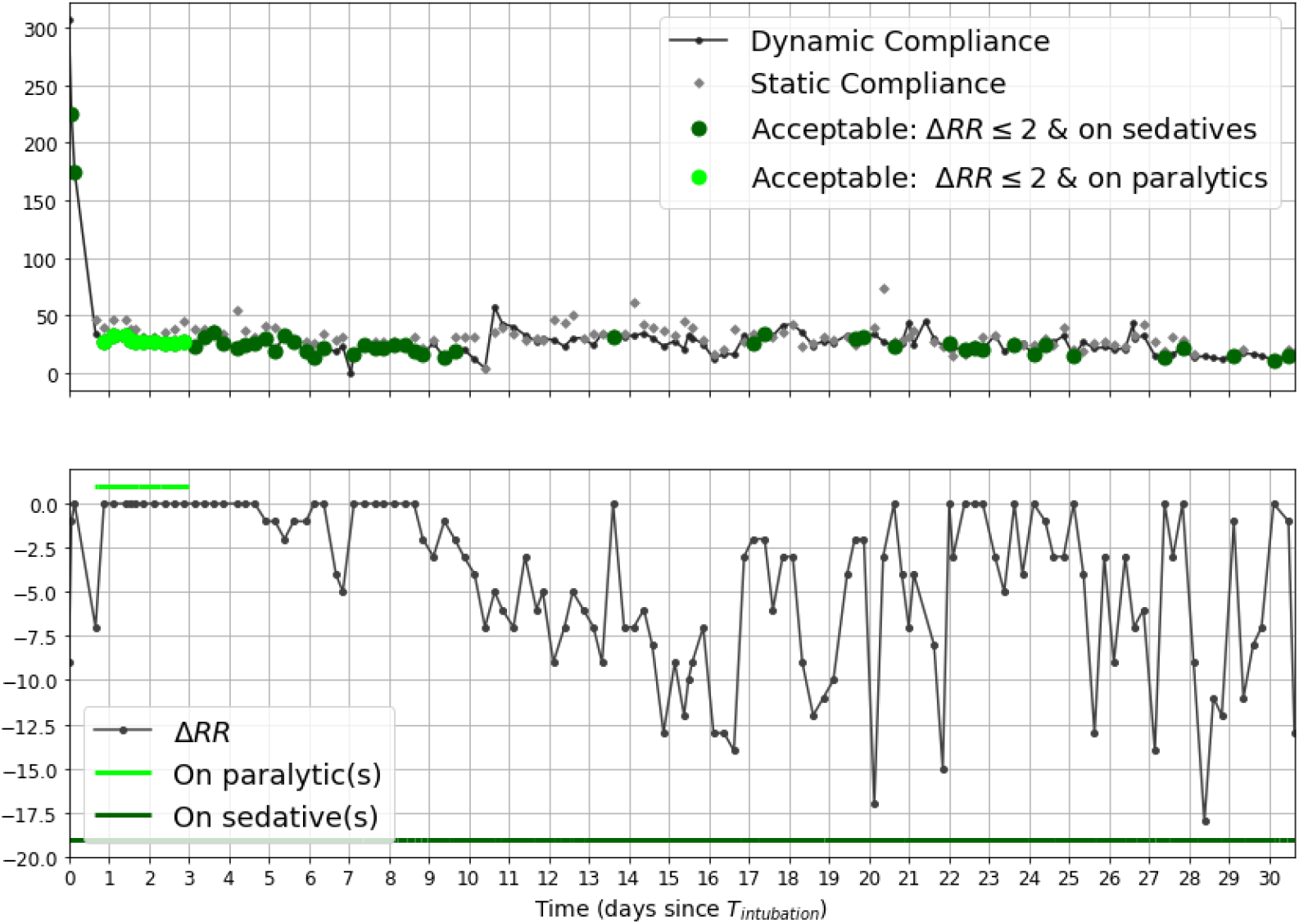
Visualization of assumptions made to detect valid respiratory system compliance measurements using a sample patient. The top figure shows all available measurements which could be used to calculate compliance, with green dots indicating when the measurements were deemed to be acceptable by the algorithm based on either being on a continuous paralytic agent or sedative with difference in set versus measured respiratory rate less than 2 breaths per minute (ΔRR <2). The bottom figure shows the ΔRR which corresponds to each timepoint of valid compliance measurement. A measurement is acceptable (green dots in the top figure) if the patient was deeply sedated and or paralyzed (green lines in the bottom figure), and were breathing at the rate set by the ventilator (ΔRR <2 in the bottom figure).

